# Higher polygenic scores for empathy increase posttraumatic stress severity in response to certain traumatic events

**DOI:** 10.1101/2021.07.26.21261139

**Authors:** Frank R Wendt, Varun Warrier, Gita A Pathak, Karestan C Koenen, Murray B Stein, John H Krystal, Robert H Pietrzak, Joel Gelernter, Elizabeth V Goldfarb, Simon Baron-Cohen, Renato Polimanti

**Author notes:** Joint first authors (Frank R Wendt, Varun Warrier). Joint senior authors (Simon Baron-Cohen, Renato Polimanti). Corresponding authors: Frank R Wendt (; & Renato Polimanti (; Department of Psychiatry, Yale University School of Medicine, VA CT 116A2, 950 Campbell Avenue, West Haven, CT 06516, USA.

## Abstract

**Background:** Posttraumatic stress disorder (PTSD) is triggered by environmental stressors. Empathy may predispose an individual to respond to life events differently if high empathizers are emotionally more sensitive to trauma. For the first time, we test this hypothesis at the genetic level.

**Methods:** We applied polygenic scoring (PGS) to investigate the shared genetics linking empathy (measured using the Empathy Quotient (EQ), a self-report measure of empathy; N=46,861) and PTSD symptom severity (measured using the 6-item PTSD Checklist 6-item (PCL-6)) in the UK Biobank (N=126,219). Follow-up analyses were performed in the context of (1) experiencing any of 16 potential traumas, (2) the total number of traumas endorsed, and (3) the context of trauma. Autism, depression, generalized anxiety, and PCL-17 PGS were included as covariates to verify the specificity of the effect.

**Results:** EQ_PGS_ associated with PCL-6 (*R*^2^=0.012%, *P*=9.35×10^−5^). This effect remained significant after accounting for autism, depression, PTSD, and anxiety PGS but was observed only in those who endorsed experiencing at least one traumatic event. EQ_PGS_ showed the strongest effect on PCL-6 (*β*=2.32, s.e.=0.762, *P*=0.002) among those who endorsed childhood neglect/abuse (*felt hated as a child*). In the highest EQ_PGS_ decile, feeling *hated as a child* was associated with lower odds of healthy adulthood interpersonal relationships (OR=0.623, 95%CI 0.443-0.885) but this association was not seen in the lowest EQ_PGS_ decile.

**Conclusions:** A genetic predisposition to higher empathy, which may index greater emotional sensitivity, predisposes an individual to more severe PTSD symptoms, specifically in the presence of early negative life events.

## INTRODUCTION

The lifetime prevalence of posttraumatic stress disorder (PTSD) is 4-7% (1). PTSD is unique among psychiatric disorders in that an environmental exposure, often termed an “index trauma,” is a core criterion for diagnosis. Genome-wide association studies (GWAS) have detected a common variant heritable component for PTSD of between 5-20% (2, 3) but we do not yet understand the underlying cognitive basis that might mediate this genetic risk of PTSD. Here we postulate that if empathy is an index of emotional sensitivity, then individual differences in empathy may predispose an individual to differential risk of PTSD.

Empathy is the ability to identify other peoples’ mental states (their thoughts, intentions, desires, and emotions), and respond to their mental states with an appropriate emotion. Higher empathy is associated with better social and communication skills (4) and greater prosocial behaviors (e.g., helping others, sharing, donating) (5). However, dysregulation of empathy can lead to greater risk for internalizing disorders, including depression (6, 7). Empathy is itself partly heritable (4, 8, 9). What is not yet known is whether a higher empathy polygenic scores (PGS) are associated with PTSD symptoms. We tested this using the Empathy Quotient (EQ), a self-report instrument that has been widely used and validated (10) and shows a partly genetic component (4).

To understand if empathy is genetically associated with PTSD symptom severity, we first tested for pair-wise genetic correlation among the traits of interest. Next, we evaluated how PGS for empathy correlates with PTSD symptoms, also considering several other PGS derived from traits associated with PTSD and/or empathy (i.e., specifically, autism, depression, and anxiety). Finally, we tested which potentially traumatic experiences affect the genetic overlap between empathy and PTSD. Among the potentially traumatic experiences investigated, we observed that the relationship between early life events and PTSD may be affected by the genetic effects of EQ among victims of childhood abuse/neglect. An overview is provided in Fig 1.

**Fig. 1.**
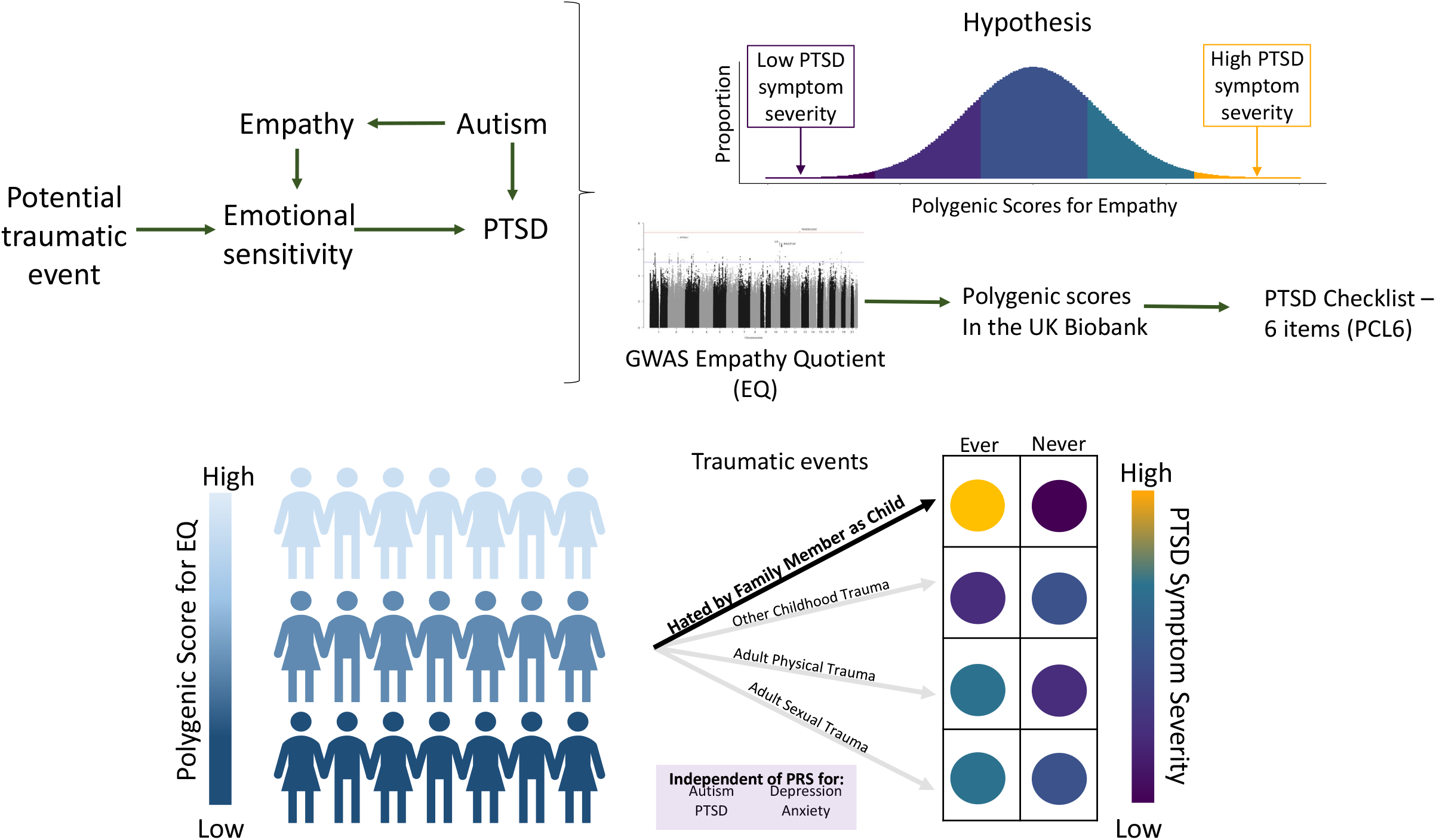
Overview of study hypothesis and analytic plan.

## METHODS

### Genome-wide Association Studies (GWAS) Statistics

The EQ GWAS consisted of participants drawn from 23andMe, Inc. A total of 46,861 participants (24,543 females and 22,318 males) completed a 60-question self-reported assessment of both cognitive and affective empathy. With high test-retest reliability (10), 40 questions were used to derive EQ per participant. EQ questions were scored from 0-2 for a maximum EQ of 80. For GWAS participants, the mean EQ was 46.4±13.7 (4).

To test if the effect of EQ on PTSD symptom severity as measured using the PTSD Checklist -6 item version (PCL-6) was independent of the relationship between polygenic scores (PGS) for autism, PTSD, generalized anxiety (GAD), and depression, all of which exist as a continuum of risk in the general population regardless of an individual’s diagnostic status, we included PGS of these phenotypes in the model of PCL-6. Autism_PGS_ was derived from a large GWAS of autism in 46,350 individuals of European descent (11). Depression_PGS_ was derived from the largest non-overlapping sample of depression phenotypes from the Psychiatric Genomics Consortium consisting of 185,720 participants (Wray, et al. association statistics with 23andMe and UK Biobank removed (12)). To avoid sample overlap between base and target datasets (UKB PCL-6 overlaps with Nievergelt, et al. (2)), we derived PTSD symptom severity PGS (PCL-17_PGS_) from the large Million Veteran Program (MVP) GWAS of the PCL 17-item questionnaire. MVP respondents (N=186,689) were asked to report the extent to which they had been affected in the previous month by symptoms in response to stressful life experiences. Each item was scored on a five-point severity scale (1=“not at all” to 5=“extremely”). GAD-2_PGS_ was derived from a large GWAS of GAD 2-item questionnaire ascertained in the MVP. Respondents (N=175,163) were asked to respond according to their symptoms during the past two weeks on a scale of 0=“not at all” to 3=“nearly every day.” As previously shown, there is a strong overlap between the GWAS of PCL-17 versus PCL-6 and GAD-2 versus ANGST and iPSYCH anxiety GWAS so demographic differences between the MVP and UKB were not expected to influence the use of these cohorts for PGS analysis (3, 13). We tested SNP-based heritability and the *r*_*g*_ between these phenotypes using Linkage Disequilibrium Score Regression (14) and the 1000 Genomes Project Phase 3 European ancestry reference panel.

### UKB Individual Level Data

The UK Biobank (UKB) is a large-scale cohort recruited across the United Kingdom to study human health and disease. Participants’ age ranged from 37-73 at the time of data collection. Following their initial visit, participants were invited to participate in an online Mental Health Questionnaire (MHQ) ultimately completed by approximately 157,000 participants. Among related individuals, we retained the individual with the higher PCL-6 score resulting in 126,219 unrelated participants of European ancestry.

PCL-6 is a summed score of six questions from the UKB MHQ (15). Participants were asked to rank the extent to which they have been affected by five PTSD symptoms in the past month (0=“Not at all” to 4=“Extremely”). PCL-6 items are UKB Field ID 20494 “felt irritable or had angry outbursts in the past month,” UKB Field ID 20495 “avoided activities or situations because of previous stressful experience in the past month,” UKB Field ID 20496 “felt distant from other people in the past month,” UKB Field ID 20497 “repeated disturbing thoughts of stressful experience in the past month,” and UKB Field ID 20498 “felt very upset when reminded of stressful experience in the past month.” The sixth item, UKB Field ID 20508 “recent trouble concentrating on things,” was answered with respect to symptoms over the last two weeks with answers ranked from 1=“Not at all” to 4=“Nearly every day” (15). The mean PCL-6 score was 6.59±3.68 (*N*=126,219) among unrelated UKB participants of European ancestry. PCL-6 scores were stratified into PTSD cases and controls using PCL-6 threshold >13 (N_case_=11,666; N_control_=114,553) (2).

### Traumatic Experience Definitions

Participants responded to 16 questions about their exposure to potentially traumatic experiences across their lifetime (UKB Category 145 Traumatic Events). Each question was ranked from 0=“never true” to 4=“very often true.” To improve power and overcome the relative rarity of several potentially traumatic experiences, we binned responses into “never” (only participants who responded with “never true”) and “ever” (participants with any exposure to the indicated potentially traumatic experience). Four items (UKB Field ID 20489: *felt loved as a child*, UKB Field ID 20491: *someone to take you to the doctor when needed as a child*, UKB Field ID 20522: *been in a confiding relationship as an adult*, and UKB Field ID 20525: *able to pay rent/mortgage as an adult*) are considered favorable experiences and were inversely coded such that higher scores indicate more frequent experiences of the potential trauma. We derived two additional variables per participant: (i) “any trauma” was coded as 0 if the participant responded “Never true” to all potential traumas (N=40,761) and 1 if the participant endorsed experiencing any frequency of any potential trauma (N=85,458) and (ii) “total number of endorsed traumas” was the summed total of all endorsed potentially traumatic experiences (mean=1.46±1.53).

### Polygenic Scoring (PGS)

PGS analysis was performed using PRSice v2 (16) with GWAS statistics as the base data to predict PCL-6 scores. GWAS data were clumped using the following parameters to identify approximately linkage disequilibrium independent SNPs: clump-*r*^2^=0.001 in 10,000-kb windows resulting in 3,680 independent EQ SNPs, 2,434 autism SNPs, and 11,137 PCL-17 SNPs, 31,967 GAD-2 SNPs, and 33,065 depression SNPs contributing the PGS calculation. The differences in the number of SNPs are due to the genotyping arrays and imputation produced used in the original studies (3, 4, 11-13). We tested ten PGS P-value thresholds (*P*_*T*_): 5×10^−8^, 1×10^−6^, 1×10^−5^, 1×10^−4^, 0.001, 0.05, 0.1, 0.3, 0.5, and 1. Stringent SNP clumping was applied at this stage to study the effects of the same collection of SNPs across several analyses (see **Mendelian Randomization**).

Enrichment analysis was conducted using PRSet implemented in PRSice v2. Gene sets were derived from the Molecular Signatures Database (MSigDB) (17) including for 5,552 Gene Ontologies (GO). Multiple testing correction was applied using a false discovery (FDR) rate of 5% to account for the correlation among the gene set annotations. Gene set analysis with PRSet was performed using *P*_*T*_=1 because it is unclear whether a gene set is associated with the phenotype when the best theshold contained only a small portion of SNPs within the gene sets, as is indeed true for EQ.

PRSice and PRSet models were performed two ways. First, we tested a baseline model of EQ_PGS_→PCL-6 with age, sex, age × sex, and the first ten principal components of ancestry as covariates:

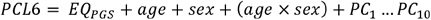

Second, we tested a full model multiple including psychopathology PGS as covariates (e.g., EQ_PGS_→PCL-6 including all baseline covariates plus autism_PGS_, depression_PGS_, GAD-2_PGS_, and PCL-17_PGS_):

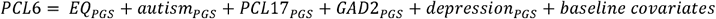

To contextualize the effect of EQ_PGS_ on PCL-6 among the traumatic experiences collected for UKB participants, the fully adjusted model was further tested among participants who did and did not endorse each event (see **Traumatic Experience Definitions)**.

### Mendelian Randomization

Mendelian randomization (MR) uses genetic instruments (i.e., SNPS) as non-modifiable exposures with which to test the causal relationship between two phenotypes (18). The PGS P_T_ producing the strongest effect between EQ and PCL-6 was used to include SNPs in the instrumental variable (19). Inverse variance weighted (IVW) estimates were generated with the `TwoSampleMR` R package testing two hypotheses: (i) EQ has a causal effect on PCL-17 and (ii) PCL-17 has a causal effect on EQ. To appropriately account for a possible weak instrument bias (genetic instrument based on variants with P-values > 5×10^−8^), we also report the robust adjusted profile score (MR-RAPS) effect size estimate. To test for effect size outliers biasing the genetic instruments in IVW or RAPS, we evaluated Cochran’s Q and its associated P-value for the hypothesis that the genetic instruments show no evidence of effect size heterogeneity.

### Modeling PTSD Diagnosis Probability

A logistic regression model of PTSD status was created using the R package `effects` (20). PTSD cases were defined as any individual with a PCL-6 > 13 (N_case_=11,666; N_control_=114,553). We sampled EQ_PGS_ 500 times per sex per endorsement of feeling hated as a child for 2,000 total samplings at fixed effects of age, total number of traumas endorsed, autism_PGS_, PCL-17_PGS_, depression_PGS_, GAD-2_PGS_, and ten within-ancestry principal components. In this way, the sampled EQ_PGS_ were solely used to model the probability of PTSD status among those who endorsed “ever” and “never” *experiencing feeling hated as a child*.

### Identifying Traumatic Event Correlates

To identify correlates of *feeling hated as a child*, we selected traits from the UKB with a *priori* support for an effect on PTSD. These included all other traumatic experiences, the specific PTSD symptoms included in the PCL-6, neuroticism score, Townsend deprivation index, income, and educational qualifications. The neuroticism score (UKB Field ID 20127) is a summary of twelve items including mood swings, fed-up feelings, nervousness, and loneliness/isolation, and other behaviors (21). The Townsend deprivation index (UKB Field ID 189) is a score of regional deprivation based on unemployment, household overcrowding, non-car ownership, and non-home ownership (22). Average total household income before tax (UKB Field ID 738) was binned into five strata ranging from *less than £18,000 to greater than £100,000*. Educational qualifications (UKB Field ID 6138) range from *other professional qualifications (e.g*., *nursing, teaching) through college or university degree*. Generalized linear models were used to test the relationship between felt hated as a child and each of the indicated variables.

## RESULTS

### Genetic Correlation

We first investigated the shared genetics between the EQ and PCL-17, GAD-2, depression, and autism. All GWAS had significant non-zero SNP-heritability estimates (Table S2). Autism and PCL-17 (*r*_*g*_ =0.342, s.e.=0.089, *P*=1.22×10^−4^) were positively genetically correlated but showed inverse *r*_*g*_ with EQ (PCL-17 *r*_*g*_ =0.117, s.e.=0.046, *P*=0.011; autism *r*_*g*_ =-0.273, s.e.=0.073, *P*=1.84×10^−4^).

### Polygenic Association of EQ and PCL-6

We next tested if PGS for empathy, autism, depression, and anxiety are associated with PCL-6 scores in the UK Biobank. After multiple testing correction for the number of *P*_*T*_ and traits tested (N=55 tests; FDR q<0.05), EQ_PGS_, autism_PGS_, depression_PGS_, GAD-2_PGS_, and PCL-17_PGS_ were all associated with greater PCL-6 scores (EQ_PGS_ *R*^2^=0.012%, *P*=9.35×10^−5^, *P*_*T*_ =1×10^−5^, autism_PGS_ *R*^2^=0.012%, *P*=1.10×10^−4^, P =1×10^−6^, depression_PGS_ *R*^2^=0.102%, *P*=1.62×10^−30^, *P*_*T*_ =0.001, GAD-2_PGS_ *R*^2^=0.098%, *P*=2.45×10^−29^, *P*_*T*_ =0.3, and PCL-17_PGS_ *R*^2^=0.050%, *P*=6.66×10^−16^, *P*_*T*_ =0.001; Fig 2 and Table S3). The effects of all PGS were independent of one another with consistent magnitudes of effect regardless of covariate combinations (Table 1). With respect to the EQ_PGS_→PCL-6 association, we identified suggestive evidence (P<0.05; Table S4) of the involvement of genes related to regulation of organelle assembly (*R*^2^=0.009%, Z=3.41, *P*=6.56×10^−4^; Table S4) and signal transduction by P53 class mediator (*R*^2^=0.009%, Z=-3.44, *P*=5.93×10^−4^).

**Table 1.** Best-fit main effects of empathizing quotient (EQ), autism, posttraumatic stress disorder 17-item questionnaire symptom count (PCL-17), depression, and generalized anxiety disorder 2-item questionnaire total score (GAD-2) polygenic scores (PGS) on PTSD Checklist 6-item summed score (PCL-6) in baseline models and models fully covaried with the inclusion of genetic load for each other psychopathology.

| Base | Model | R<sup>2</sup> (%) | P-value | P-value Threshold | P-value for effect size attenuation |
| --- | --- | --- | --- | --- | --- |
| EQ | Baseline* | 0.012 | $9.35 \times 10^{-5}$ | $1 \times 10^{-5}$ | 0.992 |
| EQ | Baseline + autism<sub>PGS</sub> + PCL-17<sub>PGS</sub> + depression<sub>PGS</sub> + GAD-2<sub>PGS</sub> | 0.011 | $9.69 \times 10^{-5}$ | $1 \times 10^{-5}$ | |
| Autism | Baseline* | 0.012 | $1.10 \times 10^{-4}$ | $1 \times 10^{-6}$ | 0.880 |
| Autism | Baseline + EQ<sub>PGS</sub> + PCL-17<sub>PGS</sub> + GAD-2<sub>PGS</sub> + depression<sub>PGS</sub> | 0.010 | $2.55 \times 10^{-4}$ | $1 \times 10^{-6}$ | |
| PCL-17 | Baseline* | 0.050 | $6.66 \times 10^{-16}$ | 0.001 | 0.332 |
| PCL-17 | Baseline + autism<sub>PGS</sub> + EQ<sub>PGS</sub> + GAD-2<sub>PGS</sub> + depression<sub>PGS</sub> | 0.034 | $2.68 \times 10^{-11}$ | 0.001 | |
| Depression | Baseline* | 0.102 | $1.62 \times 10^{-30}$ | 0.001 | 0.849 |
| Depression | Baseline + autism<sub>PGS</sub> + EQ<sub>PGS</sub> + PCL-17<sub>PGS</sub> + GAD-2<sub>PGS</sub> | 0.097 | $3.27 \times 10^{-29}$ | 0.001 | |
| GAD-2 | Baseline* | 0.098 | $2.45 \times 10^{-29}$ | 0.3 | 0.511 |
| GAD-2 | Baseline + autism<sub>PGS</sub> + EQ<sub>PGS</sub> + PCL-17<sub>PGS</sub> + depression<sub>PGS</sub> | 0.081 | $1.12 \times 10^{-24}$ | 0.3 | |

\*Baseline model covariates were age, sex, age × sex, and the first ten within-ancestry genetic principal components

**Fig. 2.**
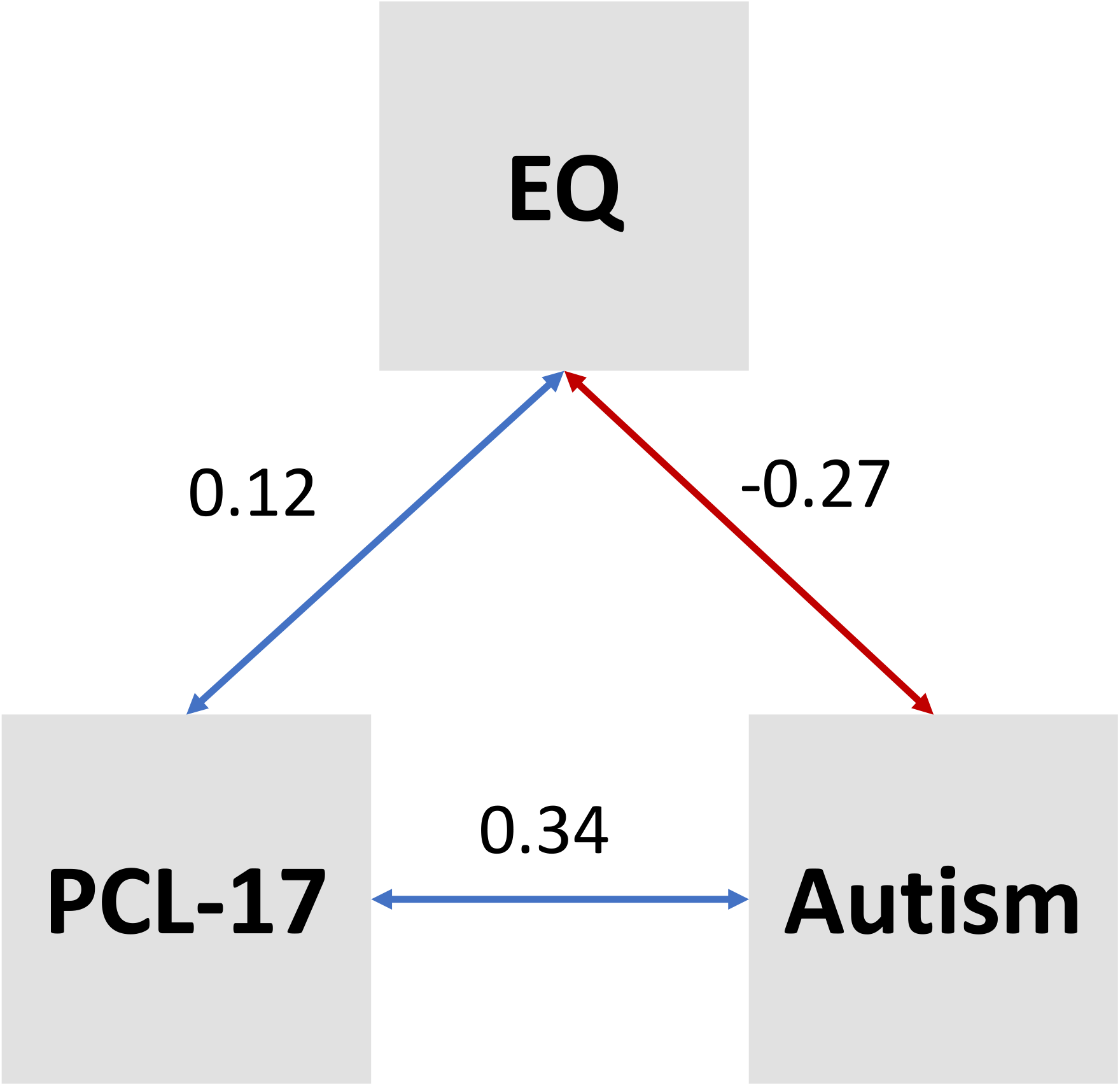
Genetic correlation (*r*_*g*_) between the PTSD Checklist 17-item symptom count (PCL-17), Empathy Quotient (EQ), and autism. Blue and red lines indicate significant positive and negative *r*_*g*_, respectively, with the magnitude of *r*_*g*_ labeled for each significant estimate.

We next applied a two-sample Mendelian randomization (MR) framework to test for the putative causal effect between EQ and PTSD using the largest available GWAS of PCL-17 (3). In the absence of heterogeneity and horizontal pleiotropy among a subset of 17 EQ SNPs (*P*_*T*_ <1×10^−5^), we detected no evidence of an effect linking EQ to PTSD symptom severity (Table S5). To ascertain whether this null result was attributed to reduced power in the EQ GWAS, we also assessed 2,200 LD-independent EQ SNPs across the full genome using MR-RAPS to correct for the possible weak instrument bias. After removing evidence of heterogeneity and horizontal pleiotropy, we also detected non-significant effect estimates between EQ and PTSD symptom severity (IVW *β*=0.005, s.e.= 0.004, *P*=0.253 and MR-RAPS *β*=0.006, s.e.= 0.005, *P*=0.236; Table S5), indicating that the relationship between EQ and PTSD may be attributed to shared biological processes/mechanisms rather than a causal effect. However, both PTSD and empathy are complex phenotypes. The GWAS of PTSD may capture behavior that increases the likelihood for experiencing traumatic events (e.g., risk taking behavior (2, 3)) and emotional sensitivity to traumatic events. We cannot distinguish between these two in the current MR analyses, and this needs to be revisited with more fine-grained phenotypes.

Furthermore, although PCL-17_PGS_ were associated nominally with EQ (*R*^2^=0.008%, *P*=0.016, *P*_*T*_=1×10^−4^), there was no evidence that genetically determined PCL-17 causally affects EQ (N=219 SNPs; IVW *β*=0.030, s.e.= 0.027, *P*=0.268 and MR-RAPS *β*=0.032, s.e.= 0.029, *P*=0.269; Table S5).

### Effect of Trauma Type on EQ and PTSD

Our primary hypothesis is that greater empathy may lead to greater emotional sensitivity, thereby contributing to PTSD symptoms after experiencing a traumatic event. To test this, the effect of EQ_PGS_ on PCL-6 was evaluated in two additional contexts: (i) in those who report ever experiencing “any trauma” versus “never trauma” and (ii) considering the total number of potentially traumatic events endorsed by each participant. In multivariable generalized linear models among participants who have experienced any trauma, the effect of EQ_PGS_ on PCL-6 (*β*=0.91, s.e.=0.281, *P*=0.001) was independent of all baseline and psychopathology PGS covariates (Table S6). There was no relationship between EQ_PGS_ and PCL-6 (*P*>0.05) in participants who report never having experienced any of the indicated potentially traumatic events, suggesting that the impact of EQ on PTSD symptoms is context specific. So, all subsequent analyses explore relationships between EQ and PTSD symptom severity in the context of total and specific traumatic experiences.

In multivariable models of PCL-6 that included the total number of traumas endorsed, autism_PGS_, GAD-2_PGS_, depression_PGS_, and PCL-17_PGS_ as covariates (Table S6), EQ_PGS_ remained associated with PCL-6 (*β*=0.569, s.e.=0.172, *P*=9.34×10^−4^). When binned by decile, participants in the highest decile of EQ_PGS_ had significantly higher PCL-6 scores (*β*=0.161, s.e.=0.057, *P*=0.005; Cohen’s *d*=0.037, 95%CI 0.013-0.062) relative to the lowest decile (Fig 3).

**Fig. 3.**
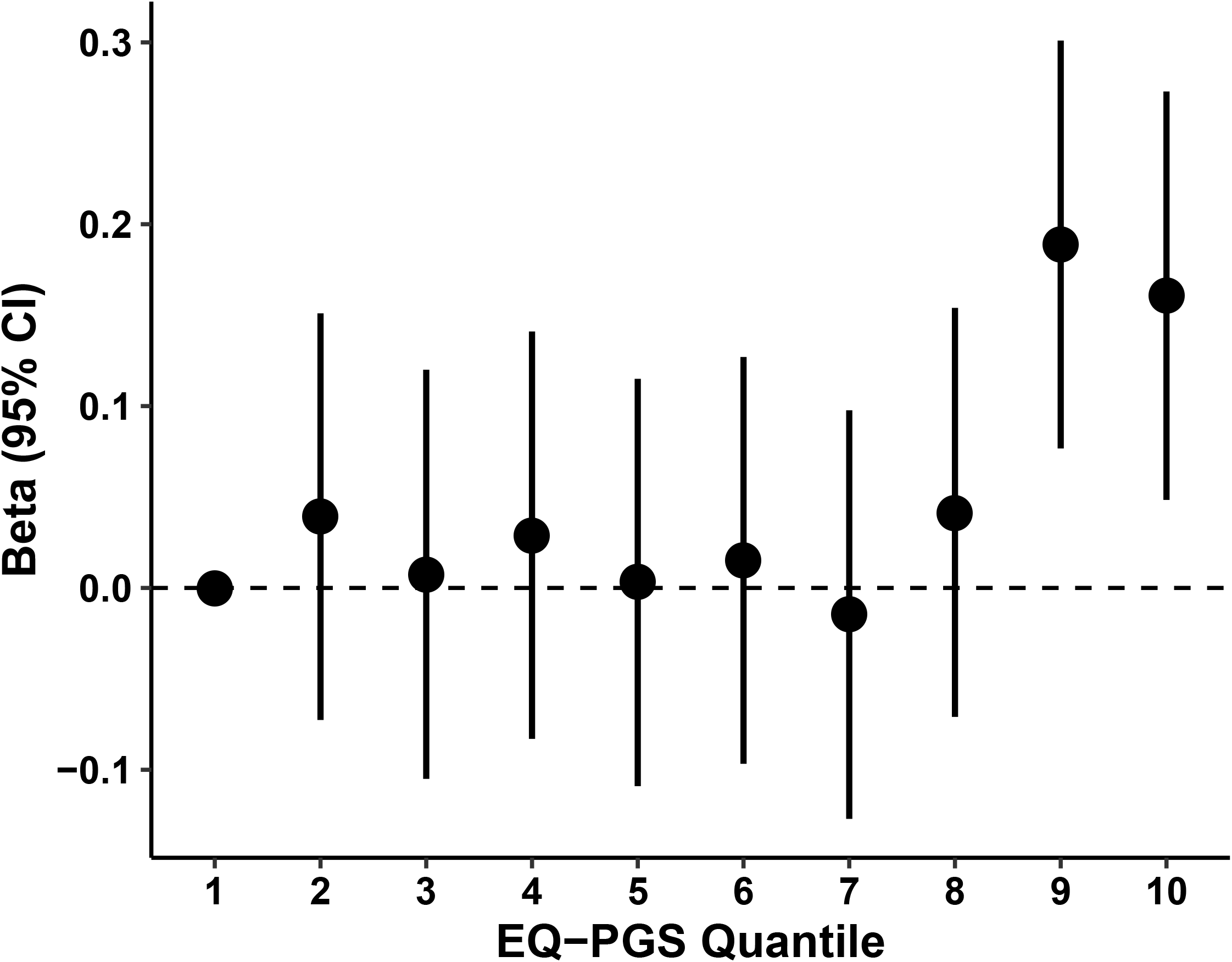
Relationship between EQ_PGS_ decile (decile 1 is the referent) and PCL-6 among UKB participants of EUR ancestry who endorsed at least one potentially traumatic experience. Effect sizes are independent of age, sex, age × sex, total number of potentially traumatic experiences endorsed, autism_PGS_, PCL-17_PGS_, depression_PGS_, GAD-2_PGS_, and ten within-ancestry principal components. Error bars represent the 95% confidence interval around each point estimate.

### Contextualized Effects of Traumatic Experiences on EQ and PTSD Relationship

To better understand if the genetic effect of empathy on PTSD symptoms differs by trauma type, we conducted PGS analyses for specific childhood and adult traumas. After multiple testing correction (FDR *q*<0.05), EQ_PGS_ associated with PCL-6 (*β*=2.32, s.e.=0.762, *P*=0.002; Table S7) most strongly among participants who “*felt hated as a child*.” This effect was independent of autism_PGS_, depression_PGS_, GAD-2_PGS_, PCL-17_PGS_, and the total number of potentially traumatic experiences endorsed (*β*=2.04, s.e.=0.727, *P*=0.005; Fig 4).

**Fig. 4.**
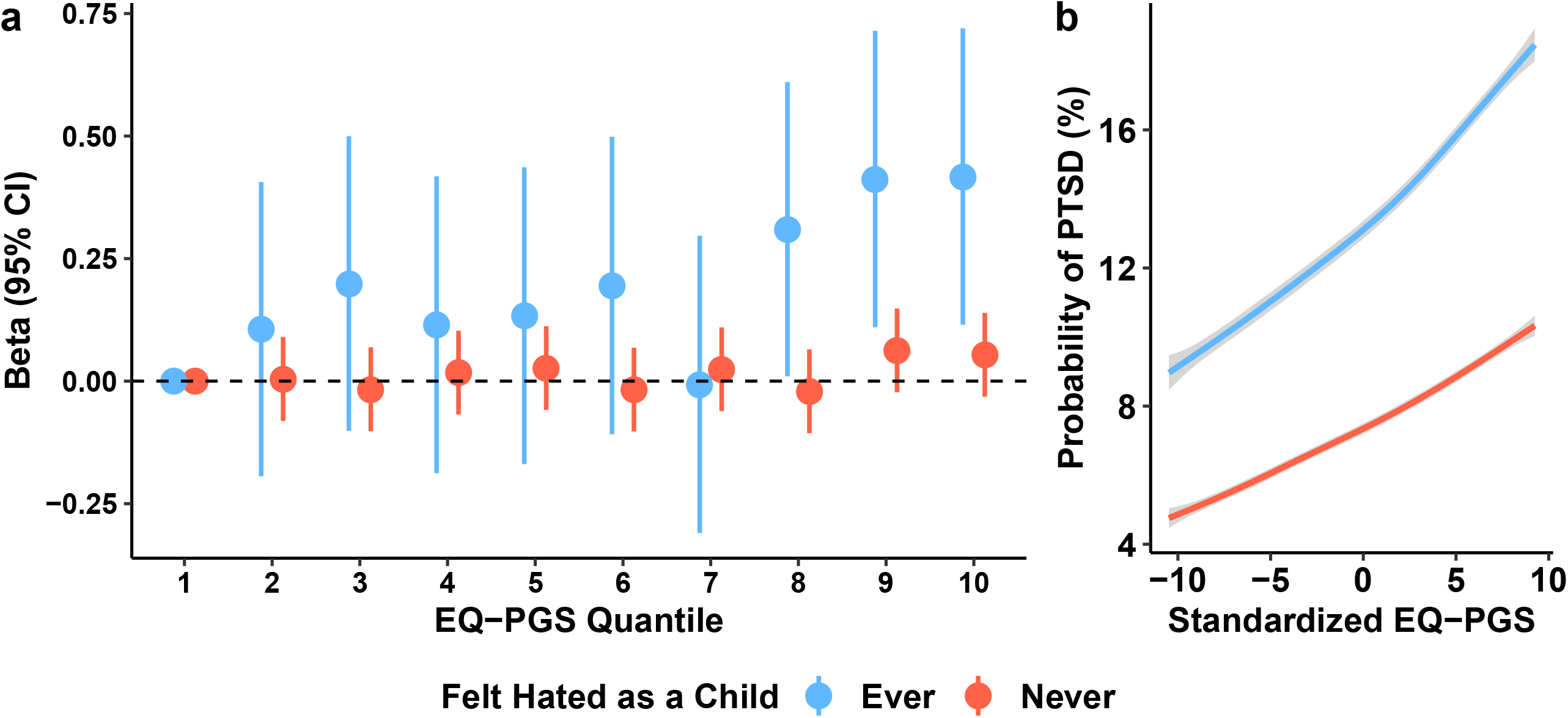
Differential effect of “*feeling hated as a child*” (“never” in blue and “ever” in red) on the relationship between empathizing quotient polygenic scores (EQ-PGS) and (a) PTSD symptom severity as measured by the PTSD Checklist 6-item questionnaire and (b) predicted probability of PTSD case-status and 95% confidence intervals (grey). All results are independent of age, sex, age × sex, total number of potentially traumatic experiences endorsed, autism_PGS_, PCL-17_PGS_, depression_PGS_, GAD-2_PGS_, and ten within-ancestry principal components. Each line in (b) represents 1,000 samplings (50% female per line) of the EQ_PGS_ at fixed covariate values.

“*Feeling hated as a child*” was endorsed by 14.5% of the UKB participants of European descent. Endorsing “*feeling hated as a child*” was associated with endorsement of “*childhood physical abuse*” (UKB Field ID 20488; OR=4.82, 95%CI 4.54-5.13, P<1.15×10^−299^), “being sexually molested as a child” (UKB Field ID 20490; OR=1.53, 95%CI 1.39-1.67, *P*=1.85×10^−19^), and “*belittlement by a partner(s) as an adult*” (UKB Field ID 20521; OR=1.53, 95%CI 1.44-1.64, *P*=3.12×10^−37^, Table S8) but none of these traumas affected the EQ_PGS_-PCL-6 association. Endorsing “feeling hated as a child” also associated with higher a Townsend deprivation index (OR=1.02, 95%CI 1.01-1.03, *P*=1.88×10^−4^) and lower educational qualifications (OR=0.96, 95%CI 0.94-0.98, *P*=2.68×10^−4^). Finally, “*feeling hated as a child*” associated with a unique pattern of current PTSD symptoms including feeling distant from others, avoidance, irritability, and repeated disturbing thoughts of the trauma (Table S8). The strongest PTSD symptom correlate of “*feeling hated as a child*” was “*feeling distant or cut-off from others in the past month*” (UKB Field ID 20496; OR=1.22, 95%CI 1.17-1.27, *P*=5.10×10^−25^). “*Feeling hated as a child*” was not associated with recent depressed or upset feelings (*P*>0.05).

When adjusted for covariates, endorsing this experience resulted in an increased probability of PTSD case-state across the spectrum of EQ polygenic score. The highest probability of PTSD was 17.93% and 10.04% for those who endorsed “*feeling hated as a child*” and those who did not, respectively (*P*_diff_=0.011; Cohen’s d=1.951, 95%CI 1.70-2.20; Fig 4 and Table S9).

Finally, we identified correlates of “*feeling hated as a child*” among the highest and lowest EQ_PGS_ deciles. Those in the highest decile of EQ_PGS_ were significantly less likely to endorse “*being in a confiding adult relationship*” (OR=0.623, 95%CI 0.443-0.885, *P*=0.007) than those in the lowest decile of EQ_PGS_ (OR=0.966, 95%CI 0.931-1.00, *P*=0.071; *P*_diff_=0.013; Table S8).

## DISCUSSION

Genetic and phenotypic overlap between psychiatric conditions and transdiagnostic traits is common and identifying their distinct and shared liabilities and possible causal relationships is crucial to understanding co-occurring conditions and long-term prognoses. Individual differences in empathy may predispose individuals to interpret specific events as more traumatic, if higher empathy entails greater emotional sensitivity (23). We hypothesized that common genetic variants underlying empathy are a risk factor for PTSD symptom severity and that this relationship may associate with specific traumatic events. As predicted, higher polygenic scores for empathy associated with more severe PTSD symptoms, particularly in individuals who report childhood neglect/abuse.

We identified independent polygenic associations of autism and EQ on PCL-6. The genetic overlap between autism and PTSD is in line with epidemiological data of high prevalence of PTSD among autistic individuals (24) and recapitulates recent genomic structural equation modeling findings. Grotzinger, et al. (25) reported autism loading onto a neurodevelopmental common factor while PTSD significantly loaded onto the same neurodevelopmental factor, as well as an internalizing factor. Interestingly, although autism and PTSD are positively correlated, EQ was negatively genetically correlated with autism but positively genetically correlated with PTSD, suggesting that EQ-associated variants may have opposite effect on autism and PTSD. All three phenotypes are complex. Specifically, with PTSD, individuals must experience a potentially traumatic event and interpret it as being traumatic (suggesting emotional sensitivity). Our results suggest that the impact of empathy on PTSD is via emotional sensitivity. There is considerable evidence indicating that autistic individuals are more likely to be maltreated by others due to lack of understanding and safeguarding (26), which increases the likelihood of experiencing traumatic events. This complex relationship between autism, PTSD, and empathy warrants further exploration.

Genetic liability to PCL-6 exists as a continuum in the general population and by stratifying the UKB into participants who did or did not endorse one of the 16 possible traumas listed in the UKB Mental Health Questionnaire, we identified the unique EQ-PCL-6 relationship in the context of those who reported exposure to abuse/neglect in childhood. Interestingly, the number of traumatic events endorsed did not attenuate the effect of EQ_PGS_ on PCL-6. When focusing on specific traumatic experiences, the EQ-PCL-6 relationship in those who endorsed experiencing childhood abuse/neglect (“*felt hated as a child*”) survived multiple testing correction and was independent of PGC for autism, depression, anxiety, and PCL-17 (27). Endorsing this life event was associated with other child abuse/neglect and belittling behavior by an intimate partner as an adult. There is evidence linking childhood maltreatment and adulthood interpersonal distress and PTSD (28, 29). In this study, participants in the highest decile of EQ_PGS_ had the lowest likelihood of having a secure confiding relationship as adults. The presence of a secure and confiding relationship in adulthood mitigates the polygenic risk for PTSD and may be a viable intervention among adult PTSD patients who report experiences of child abuse (30, 31).

Our study has several limitations. First, among the traumatic experience endorsements in the UKB, there are no indicators of self-reported severity, repeated instance measures (i.e., how many times a participant experienced an event), or indication of which trauma was the “*worst*” for each participant. While we covaried for the effect of total number of events endorsed, we could only marginally capture the full extent of this variable in the likely event that several items occur more than once across the lifespan. Second, studying the relationship between early and mid-life diagnoses requires careful consideration of age of symptom onset, age of autism diagnosis and PTSD diagnosis, and exposure to other potentially traumatic experiences across the lifespan and between diagnoses. While this study associates the genetic liability between two traits, the environmental conditions linking EQ to both autism and PTSD are likely complex. Future work in this regard will require careful identification and modeling of environmental profiles associated with autism, EQ, and PTSD liability. Third, self-reported traumatic experiences may be incomplete, potentially limited by recall bias, and limited by temporality of memory (32).

Despite these limitations, this study is the first, to our knowledge, to contextualize the relationship between empathy PGS and PTSD symptom severity among a set of potentially traumatic experiences. Consistent with prior reports of an autism-PTSD relationship (33), these findings provided additional insight into the relationship between early life events and PTSD and the mediating/moderating effect of EQ on these relationships.

## Data Availability

All data used to generate figures for this study are provided as Supplementary Material. This research has been conducted using the UK Biobank Resource (application reference no. 58146) and is available to bona fide researchers through approved access. GWAS summary statistics are available for download from several repositories: MVP (dbGAP study accession phs001672.v5.p1)and PGC.

## ACKNOWLEDGEMENTS

This research was conducted using the UK Biobank Resource (application reference no. 58146). The authors thank the research participants and employees of the UK Biobank for making this work possible. This study was supported by National Institutes of Health (R21 DC018098, R33 DA047527, and F32 MH122058) and the National Center for PTSD of the U.S. Department of Veterans Affairs. VW is funded by St. Catharine’s College, Cambridge. SBC was funded by grants from the Medical Research Council, the Wellcome Trust, the Autism Research Trust, the Templeton World Charity Foundation. SBC also received funding from the Innovative Medicines Initiative 2 Joint Undertaking (JU) under grant agreement No 777394. The JU receives support from the European Union’s Horizon 2020 research and innovation programme and EFPIA and AUTISM SPEAKS, Autistica, SFARI. His research was also supported by the National Institute of Health Research (NIHR) Applied Research Collaboration East of England (ARC EoE) programme. The funders had no role in study design, data collection and analysis, decision to publish or preparation of the manuscript.

## SUPPLEMENTARY MATERIAL

### TABLES

**Table S1**. Genome-wide association study statistics used in this study.

**Table S2**. SNP-heritability (h^2^) and genetic correlation among all genome-wide association studies in this study.

**Table S3**. Main polygenic score results between autism, Empathy Quotient (EQ), depression, and generalized anxiety disorder 2-item summed score (GAD-2) and PTSD Checklist 6-item summed score (PCL-6).

**Table S4**. Gene set enrichment results from PRSet to identify relevant gene sets contributing to the polygenic overlap between EQ and PCL-6.

**Table S5**. Two-sample Mendelian randomization testing the causal effect of EQ and PCL-17 using all SNPs contributing to the polygenic score for EQ (a), the full set of LD-independent EQ variants (b), using all SNPs contributing to the polygenic score for PCL-17 (c), and the full set of LD-independent PCL-17 variants (d).

**Table S6**. The univariable and multivariable effects of EQ_PGS_, autism_PGS_, depression_PGS_, GAD-2_PGS_, and PCL-17_PGS_ on PCL-6 in the context of broad traumatic experience reporting.

**Table S7**. The univariable and multivariable effects of EQ_PGS_ on PCL-6 in UKB participants who endorsed the given traumatic experience. The prevalence of each trauma is reported; the prevalence of Field IDs 20489, 20522, 20525, and 20491 reflect those who reported “no” as this response indicates the traumatic experience associated with these questions.

**Table S8**. Generalized linear model of endorsing “feeling hated as a child” among (a) all participants and (b) in the highest and lowest deciles of EQ_PGS_.

**Table S9**. Predicted probabilities of posttraumatic stress disorder case-status at fixed effects of all covariates (fixed at mean value per covariate). PTSD probabilities were modeled with the R package `effects` by sampling EQ_PGS_ 1,000 times per sex per endorsement of “feeling hated as a child” (2,000 total observations).

